# Prevalence and determinants of scabies: a global systematic review and meta-analysis

**DOI:** 10.1101/2024.05.06.24306963

**Authors:** Saptorshi Gupta, Simon Thornley, Arthur Morris, Gerhard Sundborn, Cameron Grant

## Abstract

**Objectives:** Scabies is a neglected skin disease that disproportionately affects people from resource poor and overcrowded countries. Global data on prevalence and risk factors are limited. This article aims to estimate the global burden of scabies and identifies the risks associated with it.

**Methods:** Databases (PubMed, Scopus and Cochrane Reviews) were accessed to identify studies of scabies prevalence published between 2000 and 2024. Results were pooled to estimate prevalence and identify factors which explained between-study heterogeneity. Odds ratios, risk of bias, subgroup analyses and meta-regression were used to describe variation in effect size and heterogeneity based on country-level demographic and economic variables.

**Results:** Seventy studies yielded a pooled prevalence of 11.9% (95% confidence interval [CI] 9.60%-14.7%) with substantial heterogeneity (1^2^ = 100%; r^2^ = 1.04). Prevalence was highest in Oceania (17.9%; 95% CI 13.9-22.8) compared to other regions. Pooled risk factors for scabies showed significant associations for demographic and behavioral factors including contact history with household members with itch (odds ratio [OR] 11.3; 95% CI 4.82-26.51; 1^2^ = 96%; *n* = 7), lack of soap use (OR 3.41; 95% CI 2.56-4.54; 1^2^ = 44%; *n* = 7), bed-sharing (OR 2.64; 95% CI 1.50-4.63; 1^2^ = 76%; *n* = 7), sharing of clothes (OR 2.52; 95% CI 1.58-4.03; 1^2^ = 85%; *n* = 7), infrequent bathing (OR 2.13; 95% CI 1.41-3.22; 1^2^ = 77%; *n* = 6), presence of pets (OR 1.76; 95% CI 1.08-2.87; 1^2^ = 84%; *n* = 4) and being a male (OR = 1.19; 95% CI 1.04-1.37; 1^2^ = 83%; *n* = 22). Socio-economic factors were not convincingly associated with scabies prevalence.

**Conclusion:** Prevalence of scabies is associated with geographic location and behavioural factors, but not between-country socioeconomic status. In addition to mass drug administration, risk factors are identified which may be included in health promotion programmes to reduce scabies prevalence and its sequelae in the long term.

## Introduction

Scabies is a parasitic and contagious disease that affects an estimated 200 million people globally, with a particularly high burden in Asia, Oceania, and Latin America (1). It leads to itchy skin, the clinical manifestations of which are characterised by small inflammatory and pruritic skin papules resembling a mosquito bite.

Scabies occurs more frequently in the young and elderly, and in immunocompromised individuals. Scabies is not benign as it is associated with complications including impetigo, cellulitis and skin abscesses, and post-infectious complications including post-streptococcal glomerulonephritis. Caused by the mite *Sarcoptes scabiei* var. *hominis,* scabies infects about 10% of children in resource-poor areas principally through human skin-to-skin contact.

Over the past decade, substantial efforts have been made by the International Alliance for the Control of Scabies (IACS) to raise the profile of scabies and prioritise control efforts (2). Recent epidemiological evidence has shown increased morbidity and mortality (3) mostly due to secondary bacterial infections occurring following scabies (2,4). Invasive secondary infections with *Streptococcus pyogenes* and *Staphylococcus aureus* (5) can lead to serious invasive bacterial infection and septicaemia. Glomerulonephritis (6) and acute rheumatic fever (7) can occur following *S. pyogenes* infection. The direct discomfort from scabies causes sleep deprivation (8), poor performance at work and reduced quality of life (9). Despite the health impacts and complications that can occur from scabies, there is limited published information about the global prevalence and distribution of the disease. Thus, to determine the prevalence and risks associated with scabies, a meta-analysis at a global scale was conducted.

## Methods

### Objectives and aim

This meta-analysis aimed to review studies describing the burden of scabies and estimate its global prevalence. We also aimed to identify factors associated with prevalence in different population settings.

### Search strategy and identification of studies

A search was conducted using PubMed, Scopus and Cochrane database. Studies published since 2000 were included. All study designs including case-control, cohort and cross-sectional studies were considered. The keywords used for search of relevant articles were ‘scabies’, ‘controlled study’, ‘major clinical study’, ‘skin disease’, ‘prevalence’, ‘skin defect’, ‘cross-sectional study’, ‘risk factor’, ‘mite’, ‘contact dermatitis’, ‘incidence’, ‘disease association’, ‘skin examination’, ‘infection’ and ‘comparative study’. The reference list of the retrieved articles was also screened for additional articles. The search strategy and number of articles reviewed at each stage is enumerated in **Supplement S1**.

### Inclusion and exclusion criteria

The exclusion and inclusion of studies were conducted according to the latest version of the PRISMA guidelines (10). Studies that clearly reported the prevalence of scabies in the form of absolute number and percentages with 95% confidence intervals in a particular country or region among a specified population were included. Studies that were freely available, published in English and could be retrieved as full-length articles were included.

Studies assessing skin conditions that did not specifically report the number of scabies cases were excluded. Additionally, articles that were published in foreign languages or were available only in subscription format were excluded.

### Outcome variable

Outcomes selected for pooled analyses were prevalence of scabies infestation and odds ratios associated with the risk of having scabies.

### Data extraction

Titles and abstract were independently reviewed by two authors (SG and ST). Subsequently studies were selected based on the inclusion and exclusion criteria. The following information was abstracted from each of the selected studies: author, year of publication, study period, prevalence (absolute number), prevalence as a percentage, method of diagnosis, country of study, World Health Organization (WHO) region, United Nations Statistics Division of countries, geographic location, population setting of the study, Gross Domestic Product (GDP) per capita in US$ of the study country, Gini index, Human Development Index (HDI), average income level of the country, age range of the population studied, the total sample size and different determinants of scabies. Among the included studies, a few were performed to determine the impact of mass drug administration on scabies prevalence (11–18). From these studies, the baseline prevalence data were used for the pooled estimates as these data describe the prevalence among the community without any intervention. The socio-economic indicators, GDP, HDI and Gini index, could only be measured at the country level (19–21).

### Quality assessment

The quality of each extracted article was assessed with the help of the Joanna Briggs Checklist for Analytical Cross-Sectional, Cohort Studies or Case-Control Studies. In studies that evaluated scabies interventions, we considered baseline prevalence only. Two reviewers (SG and ST) independently assessed risk of bias using percentage scores. A score percentage of 49 or lower was categorized as low quality whereas score percentages ranging from 50 to 69 and 70 or higher were categorized as moderate and high quality respectively. Detailed information regarding the quality of included studies is presented in **Supplementary S2**.

### Statistical analysis

The *meta*, *metafor* and *dmetar* packages of R software (version 4.3.2) (22) were used for analysis (pooling of prevalence and odds ratios), assessment (statistical tests of significance), and visualization. Results were pooled using random effects meta-analysis for primary analyses. Subgroup analyses were conducted to explore the prevalence of scabies infestation based on varying conditions like region, location, method of diagnosis, population setting and income level of the population under consideration. Meta-regression was performed on indicators of the country level socio-economic factors GDP, HDI and Gini index. Odds ratio (OR) with 95% confidence intervals (CI) were calculated separately for demographic and behavioral risk factors like contact history with household member with itch, soap use, frequency of baths, bed sharing, sharing clothes, source of water, presence of pets, location, family size, and gender.

### Assessment of Heterogeneity

The Cochran’s *Q* statistic. Higgin’s and Thompson’s 1^2^ and r^2^ statistics, with an alpha value of 0.05 were used to assess the degree of between-study heterogeneity. A *p-*value less than 0.05 for the *Q* statistic was used to confirm heterogeneity. 1^2^ was used to assess heterogeneity, as a measure of the percentage of variability in the effect size which could not be explained by sampling error (23). Values of 1^2^ equal to 50% or greater was considered ‘high’. The r^2^ statistic was used to measure the degree of between-study variance in the meta-analysis. For 1^2^ values of greater than 50%, a random effects model was used. A Baujat plot was used to detect potential outliers in the study and subsequently an influence analysis was used to check the effect of individual studies on pooled results and between-study heterogeneity on a case-by-case basis (24,25).

### Risk of Bias

The Harbord (26) and Peters test (27) were used to assess the risk of this bias in pooled odds ratios (OR). The Harbord test addresses shortcomings of the better-known Egger’s test in the case of substantial between-study heterogeneity (28). Similarly, since Egger’s test is deemed inappropriate due to its high type-I error when ORs are pooled, especially with large ORs and high between-study heterogeneity, the Peters test was used instead (27). The ‘trim and fill’ method were used to correct pooled estimates for publication bias if it was found (29).

## Results

### Description of identified studies

A total of 634 studies were identified. After removing duplicates, studies written in languages other than English, studies published before the year 2000 and non-articles, the titles, and abstracts of 502 studies were screened for relevance. Out of these, 238 studies were assessed for eligibility. In this systematic review and meta-analysis, 70 met the inclusion criteria (**Figure 1**).

**Figure 1:**
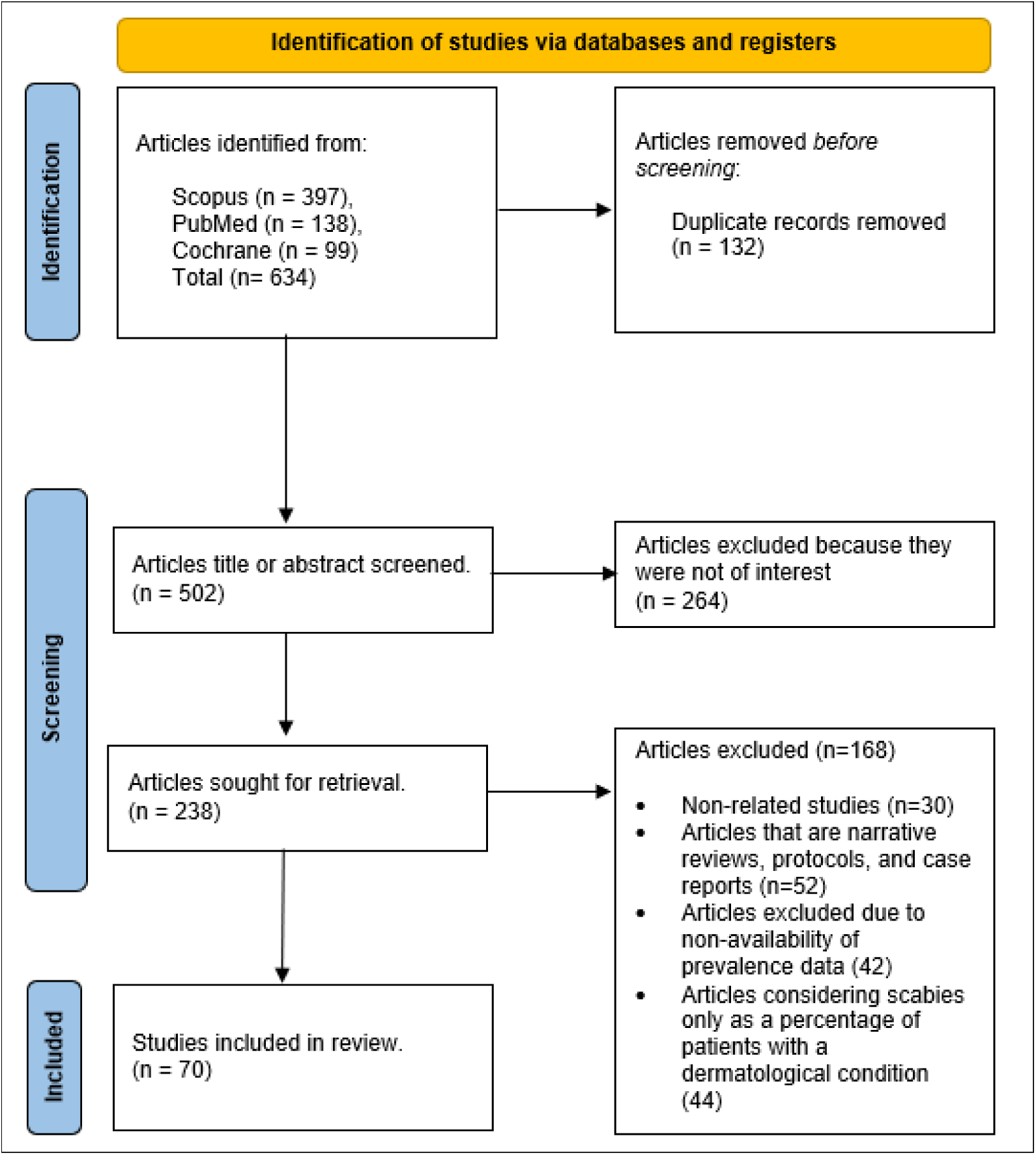
Search strategy.

All 70 included studies were conducted after 2000. The global pooled prevalence of scabies was determined from studies that included 10,324,381 participants (**Supplementary S3**). The sample size of studies ranged from 118 to 9,057,427 participants. Of the 70 eligible studies, twelve were conducted in Ethiopia (30–41), nine in Fiji (14,42–47), six in the Solomon Islands (13,16,18,48–50), four in Timor-Leste (47,51–53), five in India (11,54–57), three in Ghana (58–60) and Turkey (61–63), two in Australia (15,64), Nigeria (65,66), Malaysia (67,68), France (69,70) and Cameroon (71,72), and one each in Egypt (73), Gambia (74), Morocco (75), Iran (76), Lao (77), Liberia (78), Malawi (12), New Zealand (79), Poland (80), Guinea-Bissau (81), Botswana (82), Togo (83),Samoa (84), Bangladesh (85),Sri Lanka (86), Tanzania (17), United Kingdom (87) and Vanuatu (88). A detailed table of the study characteristics is present in **Supplementary S4**.

### Prevalence of scabies

Pooled global prevalence was 11.9% (95% CI 9.60%-14.7%). Scabies prevalence ranged widely from 0.4% to 71% (**Figure 2**), with marked variation by WHO region. Highest prevalence was recorded in the Western-Pacific region followed by the African region and the South-east Asian region. Country-level prevalence of scabies is mapped in **Figure 3**, with high prevalence in countries in the Pacific,

**Figure 2:**
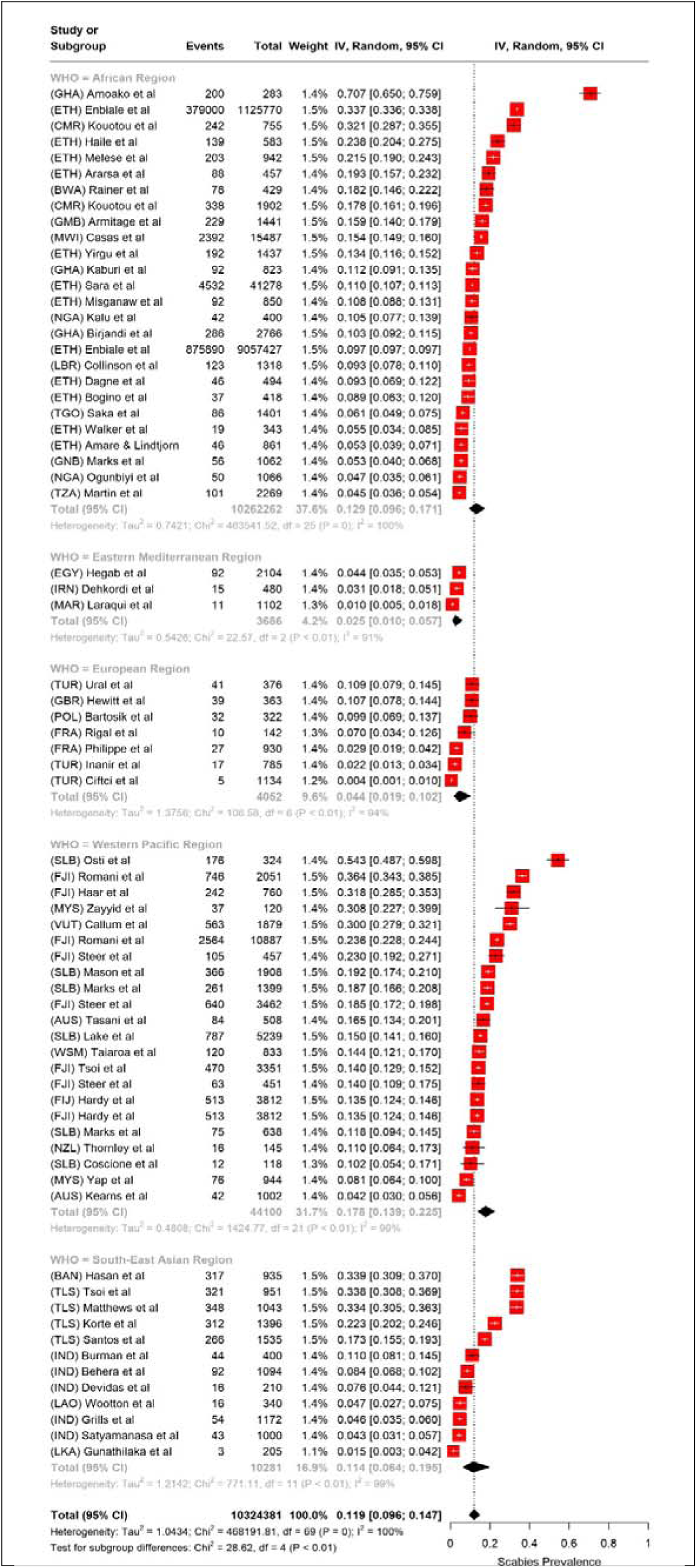
Forest plot showing pooled global scabies prevalence.

**Figure 3:**
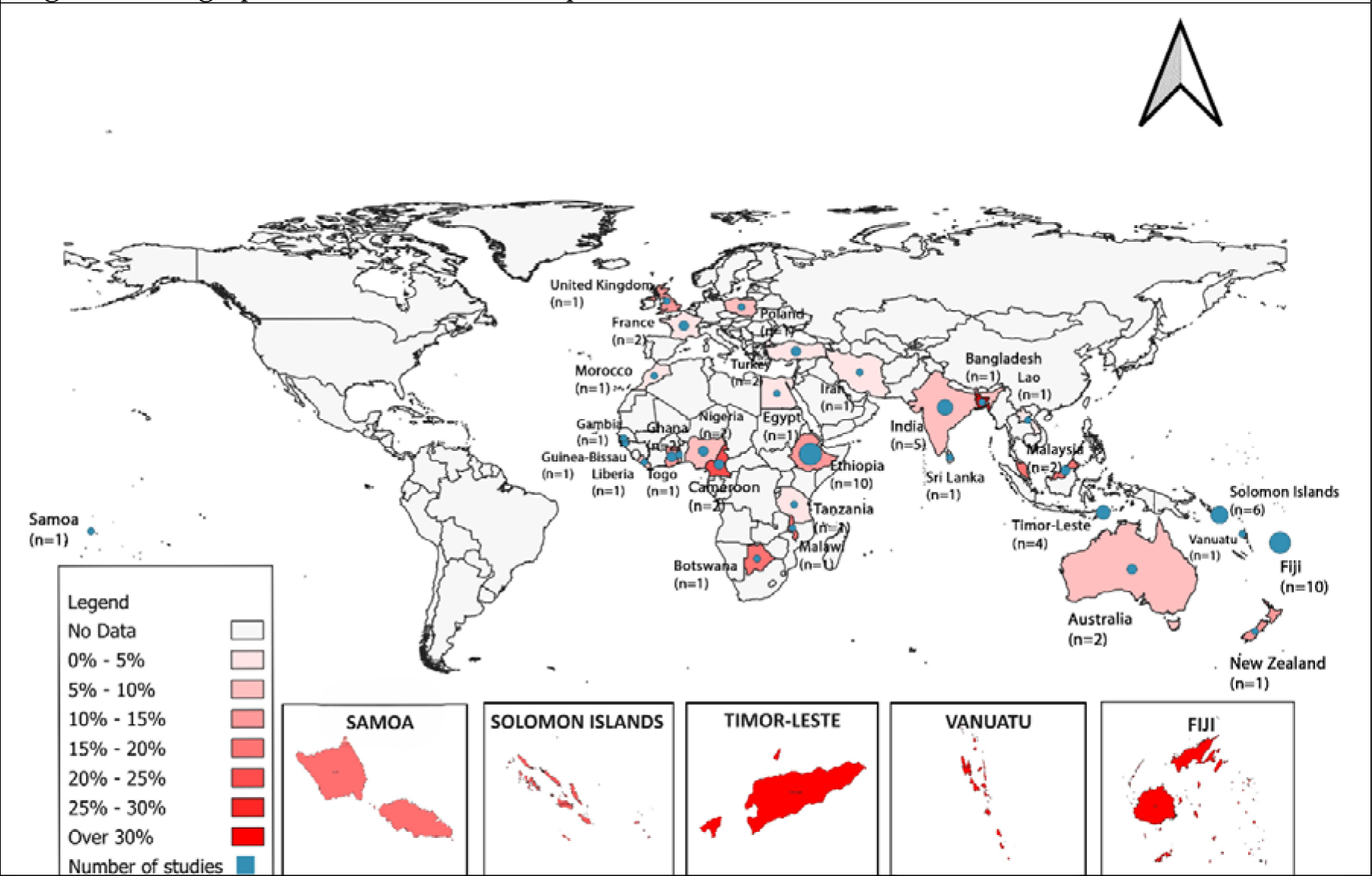
Geographic variation in scabies prevalence.

South-East Asia, and Africa. Pacific countries like Timor-Lester, Vanautu, Fiji and the Solomon Islands, marked in dark red, showed the highest pooled prevalence. Factors including different methods of diagnosis, study period, country and area under consideration explained some of the high heterogeneity in prevalence (1^2^ = 100%; r^2^ = 1.04; 95% CI: 0.75 - 1.53). Due to this high heterogeneity, a random effects model was used to compute the summary statistic. The forest plot shows the highest pooled prevalence in the Western-Pacific (17.8%; 95% CI: 13.9-22.5%), followed by Africa (12.9%; 95% CI: 9.6%-17.1%) and South-East Asia (11.4%; 95% CI: 6.4%-19.5%). The lowest prevalence region was the Eastern Mediterranean. Noticeably higher numbers of studies were published from high prevalence countries compared to low.

### Outliers and Sensitivity Analysis

The influence analysis, sensitivity analysis and Baujat plot identified four potential outliers (**Supplements S5**, **S6**, and **S7**). After removal of these studies, the pooled prevalence of scabies was 11.8% (95% CI: 9.71% to 14.26%). The sensitivity analysis also confirmed that the high heterogeneity was not dependent on a single study. Analyses hereafter have been conducted with these outliers removed.

### Subgroup Analysis

Subgroup analyses were completed after taking into consideration the method of diagnosis, WHO Region, United Regions Statistics Division (UNSD) of countries, geographic location, population settings and income status. Only region and method of diagnosis were significantly associated with prevalence. A high prevalence of 19.5% (95% CI 13.4%-27.5%) was found among studies that used the IACS criteria to diagnose scabies suggesting a higher sensitivity of the clinical definition (**Table 1**). Oceania, in the Western Pacific Region, had the highest prevalence of scabies cases. Studies that enrolled samples from specific settings, e.g., hospitals or old-age homes reported higher prevalences (13.10%; 95% CI 8.28-20.1) compared to studies performed in community settings (12.3%; 95% CI 9.63-15.6). Upper middle-income countries showed the highest pooled prevalence of scabies (15.2%; 95% CI 10.7-21.1) compared to lower-middle (12.6%; 95% CI 8.67-17.8), low (9.95%; 95% CI 7.60-12.9), and high-income countries (7.87%; 95% CI 4.96-12.3), although these differences were not statistically significant.

**Table 1:**
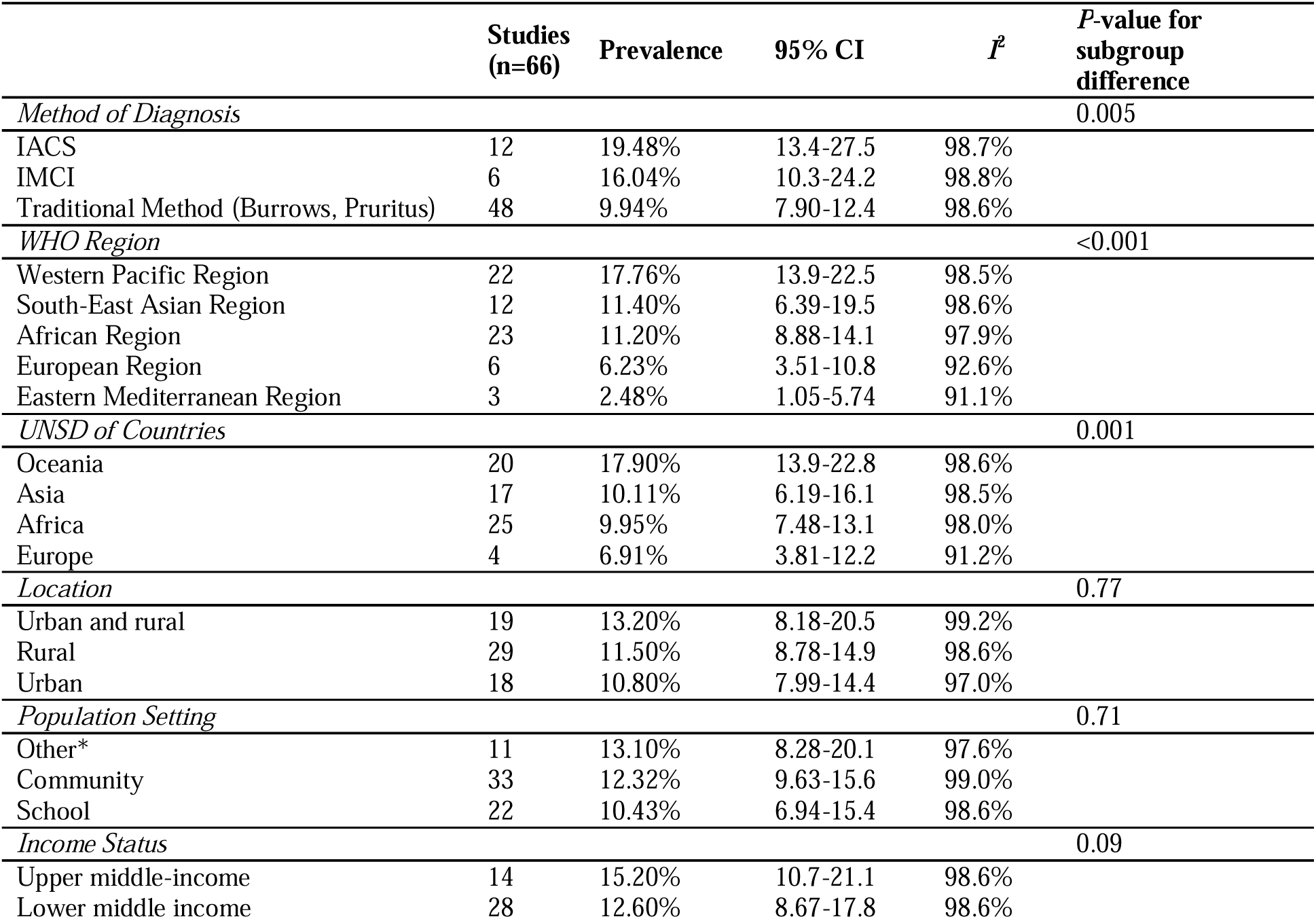
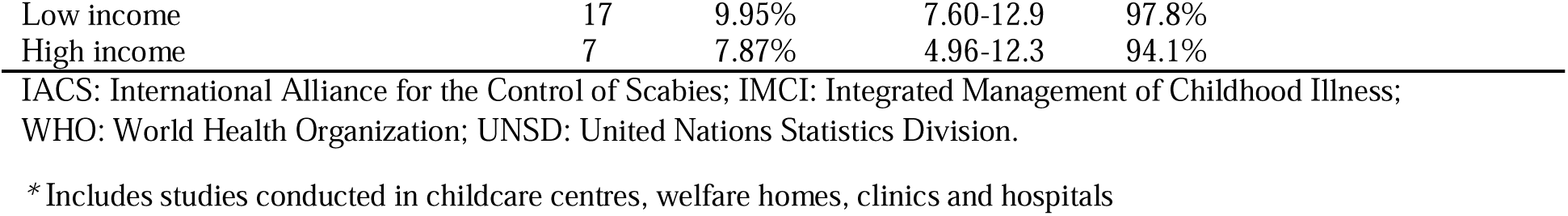
Scabies prevalence by subgroup.

| | Studies<br>(n=66) | Prevalence | 95% CI | $I^2$ | P-value for<br>subgroup<br>difference |
| --- | --- | --- | --- | --- | --- |
| *Method of Diagnosis* |  |  |  |  | 0.005 |
| IACS | 12 | 19.48% | 13.4-27.5 | 98.7% |  |
| IMCI | 6 | 16.04% | 10.3-24.2 | 98.8% |  |
| Traditional Method (Burrows, Pruritus) | 48 | 9.94% | 7.90-12.4 | 98.6% |  |
| *WHO Region* |  |  |  |  | <0.001 |
| Western Pacific Region | 22 | 17.76% | 13.9-22.5 | 98.5% |  |
| South-East Asian Region | 12 | 11.40% | 6.39-19.5 | 98.6% |  |
| African Region | 23 | 11.20% | 8.88-14.1 | 97.9% |  |
| European Region | 6 | 6.23% | 3.51-10.8 | 92.6% |  |
| Eastern Mediterranean Region | 3 | 2.48% | 1.05-5.74 | 91.1% |  |
| *UNSD of Countries* |  |  |  |  | 0.001 |
| Oceania | 20 | 17.90% | 13.9-22.8 | 98.6% |  |
| Asia | 17 | 10.11% | 6.19-16.1 | 98.5% |  |
| Africa | 25 | 9.95% | 7.48-13.1 | 98.0% |  |
| Europe | 4 | 6.91% | 3.81-12.2 | 91.2% |  |
| *Location* |  |  |  |  | 0.77 |
| Urban and rural | 19 | 13.20% | 8.18-20.5 | 99.2% |  |
| Rural | 29 | 11.50% | 8.78-14.9 | 98.6% |  |
| Urban | 18 | 10.80% | 7.99-14.4 | 97.0% |  |
| *Population Setting* |  |  |  |  | 0.71 |
| Other* | 11 | 13.10% | 8.28-20.1 | 97.6% |  |
| Community | 33 | 12.32% | 9.63-15.6 | 99.0% |  |
| School | 22 | 10.43% | 6.94-15.4 | 98.6% |  |
| *Income Status* |  |  |  |  | 0.09 |
| Upper middle-income | 14 | 15.20% | 10.7-21.1 | 98.6% |  |
| Lower middle income | 28 | 12.60% | 8.67-17.8 | 98.6% |  |

|  |  |  |  |  |
| --- | --- | --- | --- | --- |
| Low income | 17 | 9.95% | 7.60-12.9 | 97.8% |
| High income | 7 | 7.87% | 4.96-12.3 | 94.1% |

IACS: International Alliance for the Control of Scabies; IMCI: Integrated Management of Childhood Illness; WHO: World Health Organization; UNSD: United Nations Statistics Division.
\* Includes studies conducted in childcare centres, welfare homes, clinics and hospitals

### Factors associated with scabies

Separate meta-regression models showed no association of GDP, HDI or the Gini index with scabies infestation (**Supplement S8**).

### Behavioral and demographic Factors

Association of scabies status was found to be significant with the following behavioral factors – contact history with household members with itch (OR 11.3; 95% CI 4.82-26.51; 1^2^ = 96%; n=7), non-use of soap (OR 3.41; 95% CI 2.56-4.54; 1^2^ = 44%; n=7), bed sharing (OR 2.64; 95% CI 1.50-4.63; 1^2^ = 76%; n=7), cloth sharing (OR 2.52; 95% CI 1.58-4.03; 1^2^ = 85%; n=7), infrequent bathing (OR 2.13; 95% CI 1.41-3.22; 1^2^ = 77%; n=6), presence of pets (OR 1.76; 95% CI 1.08-2.87; 1^2^ = 84%; n=4) and gender (OR = 1.19; 95% CI 1.04-1.37; 1^2^ = 83%; n=22). No significant association was observed between scabies and location (OR 1.27; 95% CI 0.64-2.53; 1^2^ = 98%; n=6), use of ‘raw’ sources of water obtained from unprotected dug wells, unprotected springs, carts with small tank/drum and tanker trucks (89) (OR 1.57; 95% CI 0.60-4.06; 1^2^ = 93%; n=4), and family size (OR = 1.33; 95% CI 0.93-1.91; 1^2^ = 70%; n=7). Forest plots of these behavioral and demographic factors are illustrated in **Figure 4**.

**Figure 4:**
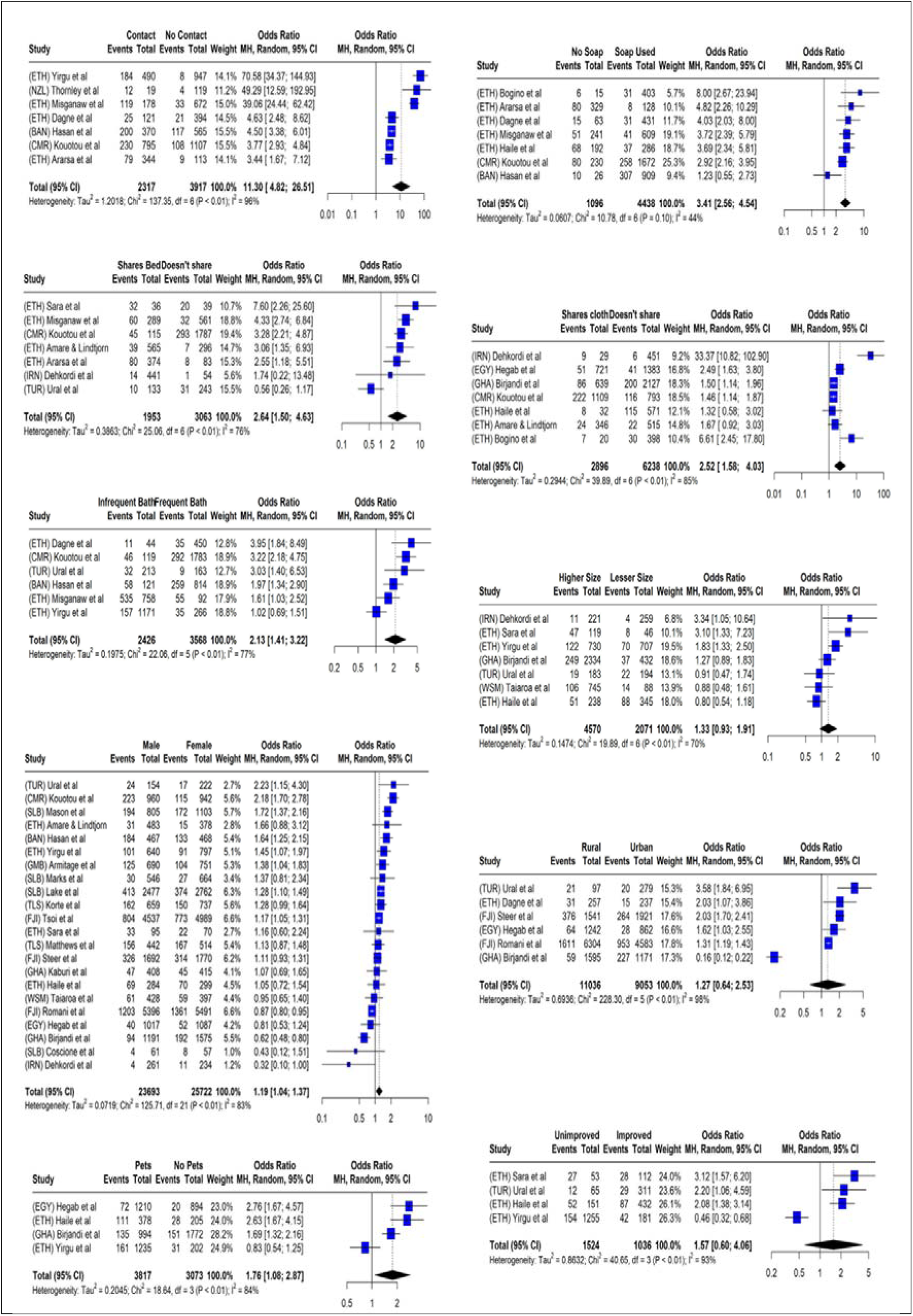
Risk factors for scabies.

### Publication bias

Meta-analyses of risk factors showed no evidence of publication bias (**Supplement S9**).

## Discussion

Our study provides a contemporary estimate of the prevalence and risk factors associated with scabies at a global level. This meta-analysis found a global prevalence of 11.9% (95% CI 9.60%-14.7%) with substantial heterogeneity (1^2^ = 100%; r^2^ = 1.04) As 1^2^ is highly dependent on the precision of the studies (90,91), it tends towards 100% as the number of studies included becomes large, since the sampling error of the pooled estimate is reduced. We found substantial heterogeneity in between-study prevalence, some of which was explained by subgroup differences. The subgroup analysis found a higher prevalence of scabies in studies conducted in the Western Pacific region. Surprisingly, country level socio-economic factors (like GDP, HDI and Gini index) were independent of scabies prevalence. Behavioral factors including infrequent bathing, lack of soap use, contact with persons with history of itch and clothes or bed-sharing were associated with higher disease risk.

A strength of this study was the comprehensive search strategy and use of PRISMA guidelines to review and include studies was likely to minimise selection bias. With 70 studies, this is the most comprehensive meta-analysis estimating the prevalence of scabies. In addition, this study considers a wide range of countries and populations with respect to geographic regions, ages, diagnostic techniques, and socioeconomic status.

This study has some limitations. First, most of the studies included were from the Western Pacific, African and South-East Asian regions, whereas only seven were from Europe and none from the Americas. This may have led to an underrepresentation and underestimation of scabies prevalence in the latter areas. Second, studies are more likely to be conducted in areas with high prevalence of scabies. Thus, pooled estimate using prevalence studies most likely overestimated the true global prevalence. Third, several risk factors such as literacy of parents and caregivers, employment status of parents, household overcrowding, and presence of sanitation facilities in the household, were not considered by the studies included in this analysis. Fourth, since different studies have considered different sets of risk factors with differing definitions, the pooled odds ratios presented in this paper is likely to be affected by measurement error. Fifth, the study has considered socio-economic factors at the country level as a proxy for every study. Since each study is based on a select population having their own unique economic characteristics, considering a macro level economic indicator may distort its association with scabies prevalence. Finally, since this study has pooled results from studies using different diagnostic techniques, this contributes to measurement error in individual studies, which likely leads to between study heterogeneity. Even though some definitions are consistent between studies, most studies rely on clinical assessment that is unavoidably subjective.

Romani et al, in 2015 (92), conducted a meta-analysis of 48 studies and concluded that scabies prevalence was highest in Latin America and Pacific regions. The findings of our study are consistent since this study also noted a significantly higher burden of scabies in the Pacific region in comparison to the others. Another recent meta-analysis (93) reported a very wide range of scabies prevalence from 0.18% to 79.6% as did the study by Romani which had a range from 0.2% to 71.4%. The pooled prevalence in our study is similar with a pooled 95% confidence interval ranging between 0.4% and 71%. We believe the main reason for such wide variation is the use of several diagnostic techniques like IACS, IMCI, and traditional methods. Thus, standardising diagnostic criteria for scabies through using the IACS method, for example, has already been recommended (94). Since these diagnostic techniques rely mainly on subjective clinical assessment, there is a need for more objective techniques such as the quantitative polymerase chain reaction (qPCR) (95). Although these meta-analyses have discussed the important issues of global prevalence, diagnostic inconsistency and need of MDA, they have, however, not considered pooling risk factors. Our study is the first to consider this evidence.

This study has considered multiple socioeconomic and behavioural factors that are likely to affect scabies prevalence. Summary measures of a country’s economic status like GDP, HDI and Gini index were not associated with scabies prevalence. This contrasts with several other ecological studies (96,97) as well as cross-sectional studies (98,99) that report evidence of higher burden of scabies in countries with lower GDP and low family income. One community-based study (98) has linked poor economic conditions to behavioural factors like infrequent bathing, soap use or washing clothes, in turn increasing the chances of scabies incidence or reinfection. The contrast in results of this study to those reported previously is likely due to the use of national GDP as a proxy for each community. There is an ecological bias in our study as summary measures at a country level don’t necessarily apply to specific populations studied. Further, since factors like income influence behaviour rather than directly influencing disease status, the role of income may be as a mediator rather than an independent causal factor.

The behavioural factors: history of contact with a person with itch; and sharing beds and clothes are consistent with past findings and are biologically plausible. Sharing clothes poses a very high risk of developing scabies (38) (99,100). Similarly, sharing towels and bed linen was strongly associated with a high prevalence of scabies cases in a study based in semi-urban India (101). Bed-sharing was strongly associated with the risk of scabies which is concordant with the results of a similar study conducted in Ethiopia (102). However, their findings of an association between family size and scabies status contrasts with those from this study. Since family size is a crude measure of crowding, it might be better to consider a measure such as household crowding index to determine the nature of this association in future studies.

In this study a strong association was observed between the presence of pets and scabies. Few studies to date have considered the risk of scabies due to pets because of the presumed biological implausibility. The pooled results from three studies provide evidence of this association. While zoonotic scabies (ZS) is often considered incapable of thriving on human skin leading to it being perceived as a self-limited disease (103), increasing evidence indicates that symptoms from ZS may persist for several weeks until an effective treatment is administered (104). Recent instances of transmission of the scabies parasite from animals, especially dogs, to humans have been reported (105,106). Also, pets may act as a ‘fomite’ which facilitates the transmission of human-to-human scabies.

The role that hygiene plays in scabies infestation is controversial and some authors dismiss these factors as not significant (107). However, several studies argue otherwise (38,98). Our study has found strong evidence of lower scabies prevalence in populations who have frequently bathe with soap. Untreated and scarce water supply leading to poor personal hygiene has been suspected to contribute to scabies burden in LMICs (38). While it is unclear if soap use directly has any preventive effect against scabies infestation, the authors of one study have stated that soap use can ameliorate the symptoms by reducing the number of active scabies lesions (108). It has also been hypothesized that regular bathing and soap use acts as a protective factor in resource poor settings (99).

Our study has provided strong evidence of the high global burden of scabies and of a number of behavioural risk factors. The evidence we have summarised indicates that specific risk factors are strongly associated with transmission. Specific measures to reduce scabies prevalence, such as mass drug administration (MDA), are now recommended by the World Health Organization for populations with a prevalence of scabies of more than 10% (96). Our study also suggests that targeted interventions that reduce risk factors such as sharing of clothes and beds and improving access to treated water and soap are also likely to reduce scabies burden.

## Conclusion

The prevalence of scabies worldwide remains high. In addition to targeted MDA, our study finds several consistent risk factors which may be included in health promotion interventions to reduce scabies prevalence and its sequelae in the long term.

## Data Availability

All data produced in the present work are contained in the manuscript

