## Supplemental File for "Prevalence and determinants of scabies: a global systematic review and meta-analysis"

**Supplementary Information**

**List of Materials**

Table S1: Search strategy

Table S2: Quality of studies

Table S3: Summary of studies used to estimate the global prevalence of scabies.

Table S4: Study characteristics

Figure S5: Leave-one out forest plot

Figure S6: Influence Analysis

Figure S7: Baujat Plot showing the outlier studies according to heterogeneity.

Table S8: Meta-regression of scabies prevalence and socioeconomic factors

Table S9: Risk of publication bias.

**Table S1: Search strategy**

**PubMed Search: 29/07/2024**

| **Search** | **Query** | **Results** |
| --- | --- | --- |
| **PubMed** | | |
| **#1** | **scabies** | **1880** |
| **#2** | **(skin diseases) OR (skin defect) OR (skin examination)** | **751,810** |
| **#3** | **(controlled study) OR (cross-sectional study) OR (major clinical study) OR (comparative study)** | **1,834,170** |
| **#4** | **(prevalence) OR (risk factor) OR (incidence) OR (disease association)** | **4,062,922** |
| **#5** | **#1 AND #2** | **1,706** |
| **#6** | **#5 AND #3** | **201** |
| **#5** | **#6 AND #4** | **138** |
| **Limit to “Human”, year 2000 to 2024, English** | | |

**Scopus**

| **#1** | **scabies** | **3,671** |
| --- | --- | --- |
| **#2** | **(skin diseases) OR (skin defect) OR (skin examination)** | **398,090** |
| **#3** | **(controlled study) OR (cross-sectional study) OR (major clinical study) OR (comparative study)** | **6,589,027** |
| **#4** | **(prevalence) OR (risk factor) OR (incidence) OR (disease association)** | **3,584,954** |
| **#5** | **#1 AND #2** | **2,045** |
| **#6** | **#5 AND #3** | **697** |
| **#5** | **#6 AND #4** | **397** |
| **Limit to “Human”, year 2000 to 2024, English** | | |

**Cochrane Search**

| **#1** | **scabies** | **325** |
| --- | --- | --- |
| **#2** | **(skin diseases) OR (skin defect) OR (skin examination)** | **33,357** |
| **#3** | **(controlled study) OR (cross-sectional study) OR (major clinical study) OR (comparative study)** | **6,589,027** |
| **#4** | **(prevalence) OR (risk factor) OR (incidence) OR (disease association)** | **1,338,480** |
| **#5** | **#1 AND #2** | **99** |
| **#6** | **#5 AND #3** | **78** |
| **#5** | **#6 AND #4** | **33** |
| **Limit to “Human”, year 2000 to 2024, English** | | |

**(((Scabies) AND ((skin diseases) OR (skin defect) OR (skin examination))) AND ((controlled study) OR (cross-sectional study) OR (major clinical study) OR (comparative study))) AND ((prevalence) OR (risk factor) OR (incidence) OR (disease association)))**

**Table S2: Quality of studies**

(a) **Analytical Cross-sectional studies**

| Author | Were the criteria for inclusion in the sample clearly defined? | Were the study subjects and the setting described in detail? | Was the exposure measured in a valid and reliable way? | Were objective, standard criteria used for measurement of the condition? | Were confounding factors identified? | Were strategies to deal with confounding factors stated? | Were the outcomes measured in a valid and reliable way? | Was appropriate statistical analysis used? | Score percentage |
| --- | --- | --- | --- | --- | --- | --- | --- | --- | --- |
| (GMB) Armitage et al | ☑ | ☑ | ☑ | ☑ | ☑ | ☑ | ☑ | ☑ | 100 |
| (CMR) Kouotou et al | ☑ | ☑ | ☑ | ☑ | ⮽ | ☑ | ☑ | ☑ | 88 |
| (EGY) Hegab et al | ⮽ | ☑ | ☑ | ☑ | ☑ | ☑ | ☑ | ☑ | 88 |
| (ETH) Ararsa et al | ☑ | ☑ | ☑ | ☑ | ⮽ | ☑ | ☑ | ☑ | 88 |
| (ETH) Haile et al | ☑ | ☑ | ☑ | ☑ | ⮽ | ☑ | ☑ | ☑ | 88 |
| (GHA) Birjandi et al | ☑ | ☑ | ☑ | ☑ | ⮽ | ☑ | ☑ | ☑ | 88 |
| (LBR) Collinson et al | ☑ | ☑ | ☑ | ☑ | ⮽ | ☑ | ☑ | ☑ | 88 |
| (VUT) Callum et al | ☑ | ☑ | ☑ | ☑ | ⮽ | ☑ | ☑ | ☑ | 88 |
| (ETH) Dagne et al | ⮽ | ☑ | ☑ | ☑ | ⮽ | ☑ | ☑ | ☑ | 75 |
| (ETH) Enbiale et al | ☑ | ☑ | ☑ | ☑ | ⮽ | ⮽ | ☑ | ☑ | 75 |
| (ETH) Melese et al | ⮽ | ☑ | ☑ | ☑ | ⮽ | ☑ | ☑ | ☑ | 75 |
| (ETH) Walker et al | ☑ | ☑ | ☑ | ☑ | ⮽ | ⮽ | ☑ | ☑ | 75 |
| (FJI) Romani et al | ⮽ | ☑ | ☑ | ☑ | ⮽ | ☑ | ☑ | ☑ | 75 |
| (IND) Satyamanasa et al | ☑ | ☑ | ☑ | ☑ | ⮽ | ⮽ | ☑ | ☑ | 75 |
| (WSM) Taiaroa et al | ☑ | ☑ | ⮽ | ☑ | ⮽ | ☑ | ☑ | ☑ | 75 |
| (SLB) Lake et al | ☑ | ☑ | ⮽ | ☑ | ⮽ | ☑ | ☑ | ☑ | 75 |
| (SLB) Mason et al | ☑ | ☑ | ⮽ | ☑ | ⮽ | ☑ | ☑ | ☑ | 75 |
| (SLB) Osti et al | ☑ | ☑ | ☑ | ☑ | ⮽ | ⮽ | ☑ | ☑ | 75 |
| (TLS) Korte et al | ⮽ | ☑ | ☑ | ☑ | ⮽ | ☑ | ☑ | ☑ | 75 |
| (TLS) Matthews et al | ☑ | ☑ | ⮽ | ☑ | ⮽ | ☑ | ☑ | ☑ | 75 |
| (GBR) Hewitt et al | ☑ | ☑ | ☑ | ☑ | ⮽ | ⮽ | ☑ | ☑ | 75 |
| (ETH) Enbiale et al | ☑ | ☑ | ⮽ | ☑ | ⮽ | ⮽ | ☑ | ☑ | 63 |
| (ETH) Misganaw et al | ☑ | ☑ | ☑ | ⮽ | ⮽ | ☑ | ⮽ | ☑ | 63 |
| (FJI) Romani et al | ⮽ | ☑ | ⮽ | ☑ | ⮽ | ☑ | ☑ | ☑ | 63 |
| (FJI) Steer et al | ☑ | ☑ | ⮽ | ☑ | ⮽ | ⮽ | ☑ | ☑ | 63 |
| (GHA) Amoako et al | ⮽ | ☑ | ☑ | ☑ | ⮽ | ⮽ | ☑ | ☑ | 63 |
| (IRN) Dehkordi et al | ⮽ | ☑ | ☑ | ☑ | ⮽ | ⮽ | ☑ | ☑ | 63 |
| (MWI) Casas et al | ☑ | ☑ | ⮽ | ☑ | ⮽ | ⮽ | ☑ | ☑ | 63 |
| (NGA) Kalu et al | ☑ | ☑ | ⮽ | ☑ | ⮽ | ⮽ | ☑ | ☑ | 63 |
| (LKA) Gunathilaka et al | ⮽ | ☑ | ☑ | ☑ | ⮽ | ⮽ | ☑ | ☑ | 63 |
| (TUR) Ural et al | ⮽ | ☑ | ☑ | ☑ | ⮽ | ⮽ | ☑ | ☑ | 63 |
| (FJI) Steer et al | ⮽ | ☑ | ⮽ | ☑ | ⮽ | ⮽ | ☑ | ☑ | 50 |
| (FJI) Tsoi et al | ⮽ | ☑ | ⮽ | ☑ | ⮽ | ⮽ | ☑ | ☑ | 50 |
| (GHA) Kaburi et al | ⮽ | ☑ | ⮽ | ☑ | ⮽ | ⮽ | ☑ | ☑ | 50 |
| (IND) Devidas et al | ☑ | ☑ | ⮽ | ⮽ | ⮽ | ⮽ | ☑ | ☑ | 50 |
| (LAO) Wootton et al | ⮽ | ☑ | ⮽ | ☑ | ⮽ | ⮽ | ☑ | ☑ | 50 |
| (MYS) Zayyid et al | ⮽ | ☑ | ⮽ | ☑ | ⮽ | ⮽ | ☑ | ☑ | 50 |
| (NZL) Thornley et al | ⮽ | ☑ | ⮽ | ☑ | ⮽ | ⮽ | ☑ | ☑ | 50 |
| (TLS) Santos et al | ⮽ | ☑ | ⮽ | ☑ | ⮽ | ⮽ | ☑ | ☑ | 50 |
| (TLS) Tsoi et al | ⮽ | ☑ | ⮽ | ☑ | ⮽ | ⮽ | ☑ | ☑ | 50 |
| (IND) Burman et al | ☑ | ☑ | ⮽ | ⮽ | ⮽ | ⮽ | ☑ | ☑ | 50 |
| (MYS) Yap et al | ⮽ | ☑ | ⮽ | ⮽ | ⮽ | ⮽ | ☑ | ☑ | 38 |
| (NGA) Ogunbiyi et al | ⮽ | ☑ | ⮽ | ☑ | ⮽ | ⮽ | ☑ | ☑ | 50 |
| (AUS) Kearns et al | ☑ | ☑ | ☑ | ☑ | ⮽ | ⮽ | ☑ | ☑ | 75 |
| (AUS) Tasani et al | ☑ | ☑ | ☑ | ☑ | ⮽ | ⮽ | ☑ | ☑ | 75 |
| (FJI) Haar et al | ☑ | ☑ | ☑ | ☑ | ☑ | ⮽ | ☑ | ☑ | 88 |
| (FJI) Hardy et al | ☑ | ☑ | ☑ | ☑ | ⮽ | ☑ | ☑ | ☑ | 88 |
| (IND) Behera et al | ☑ | ☑ | ☑ | ☑ | ⮽ | ☑ | ☑ | ☑ | 88 |
| (SLB) Coscione et al | ☑ | ☑ | ☑ | ☑ | ☑ | ☑ | ☑ | ☑ | 100 |
| (SLB) Marks et al | ☑ | ☑ | ☑ | ☑ | ⮽ | ⮽ | ☑ | ☑ | 75 |
| (SLB) Marks et al | ☑ | ☑ | ☑ | ☑ | ⮽ | ☑ | ☑ | ☑ | 88 |
| (TZA) Martin et al | ⮽ | ☑ | ⮽ | ☑ | ⮽ | ⮽ | ☑ | ☑ | 50 |

(b) **Case-Control Study**

| Study | Were the groups comparable other than the presence of disease in cases or the absence of disease in controls? | Were cases and controls matched appropriately? | Were the same criteria used for identification of cases and controls? | Was exposure measured in a standard, valid and reliable way? | Was exposure measured in the same way for cases and controls? | Were confounding factors identified? | Were strategies to deal with confounding factors stated? | Were outcomes assessed in a standard, valid and reliable way for cases and controls? | Was the exposure period of interest long enough to be meaningful? | Was appropriate statistical analysis used? | Score Percentage |
| --- | --- | --- | --- | --- | --- | --- | --- | --- | --- | --- | --- |
| (ETH) Sara et al | ☑ | ⮽ | ☑ | ☑ | ☑ | ☑ | ☑ | ☑ | ⮽ | ☑ | 80 |

(c) **Cohort Study**

| Studies | Were the two groups similar and recruited from the same population? | Were the exposures measured similarly to assign people to both exposed and unexposed groups? | Was the exposure measured in a valid and reliable way? | Were confounding factors identified? | Were strategies to deal with confounding factors stated? | Were the groups/participants free of the outcome at the start of the study (or at the moment of exposure)? | Were the outcomes measured in a valid and reliable way? | Was the follow up time reported and sufficient to be long enough for outcomes to occur? | Was follow up complete, and if not, were the reasons to loss to follow up described and explored? | Were strategies to address incomplete follow up utilized? | Was appropriate statistical analysis used? | Score percentage |
| --- | --- | --- | --- | --- | --- | --- | --- | --- | --- | --- | --- | --- |
| (FJI) Steer et al | ☑ | ☑ | ⮽ | ⮽ | ⮽ | ⮽ | ☑ | ☑ | ☑ | ⮽ | ☑ | 55 |

**Table S3: Summary of studies used to estimate the global prevalence of scabies**

| Study Characteristics | Number or Percentage | 95% C.I. Lower | 95% C.I. Upper |
| --- | --- | --- | --- |
| Number of studies | 70 |  |  |
| Number of subjects | 10,324,381 |  |  |
| Number of scabies cases | 1,275,191 |  |  |
| Prevalence of scabies | 11.9% | 9.60% | 14.7% |
| Tau-squared *(τ*^2^*)* | 1.04 | 0.74 | 1.52 |
| *I*^2^ | 100% |  |  |
| *H-*statistic | 82.4 |  |  |

C. I., confidence interval.

**Table S4: Study characteristics**

| Sl No | Author | Year of Publication | Country | Prevalence (in %) | Prevalence (absolute number) | Sample (n) | Study Period | Diagnosis | GINI | HDI | WHO | UNSD | Area | Population | GDP (per capita USD) | Income | Age Group |
| --- | --- | --- | --- | --- | --- | --- | --- | --- | --- | --- | --- | --- | --- | --- | --- | --- | --- |
| 1 | (TUR) Ciftci et al | 2006 | Turkey | 0.4 | 5 | 1134 | 2004-2005 | Traditional (Burrows, Pruritus) | 43.5 | 0.696 | European Region | Asia | Urban | School | 6031.8 | Upper Middle Income | 4-6 years |
| 2 | (MAR) Laraqui et al | 2018 | Morocco | 1 | 11 | 1102 | 2017 | Traditional (Burrows, Pruritus) | 39.5 | 0.671 | Eastern Mediterranean Region | Africa | Urban and Rural | Community | 3288.5 | Lower Middle Income | All ages |
| 3 | (LKA) Gunathilaka et al | 2019 | Sri Lanka | 1.5 | 3 | 205 | 2016-2017 | Traditional (Burrows, Pruritus) | 39.3 | 0.771 | South-East Asian Region | Asia | Urban and Rural | School | 4401 | Lower Middle Income | 5-16 years |
| 4 | (TUR) Inanir et al | 2002 | Turkey | 2.2 | 17 | 785 |  | Traditional (Burrows, Pruritus) | 43.5 | 0.685 | European Region | Asia | Rural | School | 3640.8 | Upper Middle Income | 6-14 years |
| 5 | (FRA) Philippe et al | 2005 | France | 2.9 | 27 | 930 | 2000-2003 | Traditional (Burrows, Pruritus) | 32.6 | 0.844 | European Region | Europe | Urban | Other (Childcare centre, Welfare Home, Clinic/Hospital) | 22416 | High Income | All ages |
| 6 | (IRN) Dehkordi et al | 2021 | Iran | 3.1 | 15 | 480 | 2018 | Traditional (Burrows, Pruritus) | 42 | 0.787 | Eastern Mediterranean Region | Asia | Urban and Rural | School | 3874 | Lower Middle Income | 6-13 years |
| 7 | (AUS) Kearns et al | 2015 | Australia | 4.2 | 42 | 1002 | 2010-2011 | Traditional (Burrows, Pruritus) | 34.7 | 0.923 | Western Pacific Region | Oceania | Rural | Community | 52133 | High Income | All ages |
| 8 | (IND) Satyamanasa et al | 2021 | India | 4.3 | 43 | 1000 | 2021 | Traditional (Burrows, Pruritus) | 34.2 | 0.633 | South-East Asian Region | Asia | Urban | School | 2238 | Lower Middle Income | 5-15 years |
| 9 | (EGY) Hegab et al | 2015 | Egypt | 4.4 | 92 | 2104 | 2014 | Traditional (Burrows, Pruritus) | 31.8 | 0.699 | Eastern Mediterranean Region | Africa | Urban and Rural | School | 3197 | Lower Middle Income | 6-12 years |
| 10 | (TZA) Martin et al | 2018 | Tanzania | 4.4 | 101 | 2269 | 2012 | Traditional (Burrows, Pruritus) | 37.8 | 0.504 | African Region | Africa | Rural | Community | 856 | Low Income | All ages |
| 11 | (IND) Grills et al | 2012 | India | 4.6 | 54 | 1172 | 2010 | Traditional (Burrows, Pruritus) | 35.4 | 0.572 | South-East Asian Region | Asia | Rural | Community | 1350.6 | Lower Middle Income | All ages |
| 12 | (LAO) Wootton et al | 2018 | Lao | 4.7 | 16 | 340 | 2017 | Traditional (Burrows, Pruritus) | 38.8 | 0.607 | South-East Asian Region | Asia | Rural | Community | 2440 | Lower Middle Income | All ages |
| 13 | (NGA) Ogunbiyi et al | 2005 | Nigeria | 4.7 | 50 | 1066 | 2005 | Traditional (Burrows, Pruritus) | 40.1 | 0.469 | African Region | Africa | Urban | School | 1250 | Low Income | 4-15 years |
| 14 | (ETH) Amare & Lindtjorn | 2021 | Ethiopia | 5.3 | 46 | 861 | 2017 | Traditional (Burrows, Pruritus) | 35 | 0.472 | African Region | Africa | Rural | School | 755.8 | Low Income | 7-14 years |
| 15 | (GNB) Marks et al | 2019 | Guinea-Bissau | 5.3 | 56 | 1062 | 2018 | Traditional (Burrows, Pruritus) | 34.8 | 0.48 | African Region | Africa | Rural | Community | 807.361 | Low Income | 1-9 years |
| 16 | (ETH) Walker et al | 2017 | Ethiopia | 5.5 | 19 | 343 | 2016 | Traditional (Burrows, Pruritus) | 35 | 0.47 | African Region | Africa | Rural | School | 706 | Low Income | <17 years |
| 17 | (TGO) Saka et al | 2022 | Togo | 6.1 | 86 | 1401 | 2021 | Traditional (Burrows, Pruritus) | 37.9 | 0.539 | African Region | Africa | Rural | Community | 965 | Low Income | All ages |
| 18 | (FRA) Rigal et al | 2016 | France | 7 | 10 | 142 | 2009-2011 | Traditional (Burrows, Pruritus) | 32.7 | 0.875 | European Region | Europe | Urban | Other (Childcare centre, Welfare Home, Clinic/Hospital) | 41737 | High Income | 0-11 years |
| 19 | (IND) Devidas et al | 2021 | India | 7.61 | 16 | 210 | 2015 | Traditional (Burrows, Pruritus) | 35.7 | 0.629 | South-East Asian Region | Asia | Rural | Community | 1590 | Lower Middle Income | <5 years |
| 20 | (MYS) Yap et al | 2010 | Malaysia | 8.1 | 76 | 944 | 2009 | Traditional (Burrows, Pruritus) | 45.5 | 0.762 | Western Pacific Region | Asia | Urban | School | 7167 | Upper Middle Income | 13-17 years |
| 21 | (IND) Behera et al | 2021 | India | 8.40 | 92 | 1094 | 2017-2018 | Intergrated Management of Childhood Illeness (IMCI) | 35.7 | 0.645 | South-East Asian Region | Asia | Rural | Community | 1974 | Lower Middle Income | All ages |
| 22 | (ETH) Bogino et al | 2023 | Ethiopia | 8.9 | 37 | 418 | 2020 | IACS | 35 | 0.489 | African Region | Africa | Urban | Other (Childcare centre, Welfare Home, Clinic/Hospital) | 918.7 | Low Income | 17-60 years |
| 23 | (ETH) Dagne et al | 2019 | Ethiopia | 9.3 | 46 | 494 | 2018 | Traditional (Burrows, Pruritus) | 35 | 0.489 | African Region | Africa | Urban and Rural | School | 758 | Low Income | 5-19 years |
| 24 | (LBR) Collinson et al | 2020 | Liberia | 9.3 | 123 | 1318 | 2020 | IACS | 35.3 | 0.48 | African Region | Africa | Urban | Community | 598 | Low Income | All ages |
| 25 | (ETH) Enbiale et al | 2020 | Ethiopia | 9.7 | 875890 | 9057427 | 2018 | Traditional (Burrows, Pruritus) | 35 | 0.489 | African Region | Africa | Urban and Rural | Community | 758 | Low Income | All ages |
| 26 | (POL) Bartosik et al | 2023 | Poland | 9.9 | 32 | 322 | 2014 | Traditional (Burrows, Pruritus) | 32.8 | 0.867 | European Region | Europe | Urban | Other (Childcare centre, Welfare Home, Clinic/Hospital) | 14182 | High Income | All ages |
| 27 | (SLB) Coscione et al | 2018 | Solomon Islands | 10.2 | 12 | 118 | 2018 | Intergrated Management of Childhood Illeness (IMCI) | 37.1 | 0.566 | Western Pacific Region | Oceania | Rural | Community | 2450 | Lower Middle Income | All ages |
| 28 | (GHA) Birjandi et al | 2019 | Ghana | 10.3 | 286 | 2766 | 2018 | Traditional (Burrows, Pruritus) | 43.5 | 0.62 | African Region | Africa | Urban | School | 2180 | Lower Middle Income | 11-18 years |
| 29 | (NGA) Kalu et al | 2015 | Nigeria | 10.5 | 42 | 400 | 2012-13 | Traditional (Burrows, Pruritus) | 35.5 | 0.506 | African Region | Africa | Rural | School | 2976.8 | Lower Middle Income | 6-12 years |
| 30 | (GBR) Hewitt et al | 2014 | United Kingdom | 10.70 | 39 | 363 | 2012-2013 | Traditional (Burrows, Pruritus) | 32.7 | 0.922 | European Region | Europe | Urban | Other (Childcare centre, Welfare Home, Clinic/Hospital) | 47447 | High Income | >75 |
| 31 | (ETH) Misganaw et al | 2022 | Ethiopia | 10.82 | 92 | 850 | 2020 | Traditional (Burrows, Pruritus) | 35 | 0.498 | African Region | Africa | Urban | Community | 918 | Low Income | 5-14 years |
| 32 | (TUR) Ural et al | 2022 | Turkey | 10.90 | 41 | 376 | 2021 | IACS | 41.9 | 0.838 | European Region | Asia | Urban and Rural | Other (Childcare centre, Welfare Home, Clinic/Hospital) | 9661.2 | Upper Middle Income | All ages |
| 33 | (ETH) Sara et al | 2018 | Ethiopia | 11 | 4532 | 41278 | 2016 | Traditional (Burrows, Pruritus) | 35 | 0.47 | African Region | Africa | Urban and Rural | Community | 705 | Low Income | All ages |
| 34 | (IND) Burman et al | 2020 | India | 11.00 | 44 | 400 | 2018-2019 | Traditional (Burrows, Pruritus) | 35 | 0.645 | South-East Asian Region | Asia | Urban and Rural | School | 1974 | Lower Middle Income | 13-15 years |
| 35 | (NZL) Thornley et al | 2023 | New Zealand | 11 | 16 | 145 | 2021-2022 | IACS | 32.5 | 0.937 | Western Pacific Region | Oceania | Urban | Other (Childcare centre, Welfare Home, Clinic/Hospital) | 48,781 | High Income | <14 years |
| 36 | (GHA) Kaburi et al | 2019 | Ghana | 11.20 | 92 | 823 | 2017 | Traditional (Burrows, Pruritus) | 43.5 | 0.616 | African Region | Africa | Urban | School | 1999 | Lower Middle Income | 2-7 years |
| 37 | (SLB) Marks et al | 2019 | Solomon Islands | 11.8 | 75 | 638 | 2016-2017 | Traditional (Burrows, Pruritus) | 37.1 | 0.564 | Western Pacific Region | Oceania | Rural | Community | 2283 | Lower Middle Income | All ages |
| 38 | (ETH) Yirgu et al | 2023 | Ethiopia | 13.4 | 192 | 1437 | 2018 | Traditional (Burrows, Pruritus) | 35 | 0.479 | African Region | Africa | Rural | Community | 758.3 | Low Income | All ages |
| 39 | (FIJ) Hardy et al | 2021 | Fiji | 13.5 | 513 | 3812 | 2017 | Traditional (Burrows, Pruritus) | 30.7 | 0.728 | Western Pacific Region | Oceania | Rural | Community | 5825 | Upper Middle Income | All ages |
| 40 | (FJI) Hardy et al | 2020 | Fiji | 13.5 | 513 | 3812 | 2019 | Traditional (Burrows, Pruritus) | 30.7 | 0.746 | Western Pacific Region | Oceania | Rural | Community | 5968 | Upper Middle Income | All ages |
| 41 | (FJI) Steer et al | 2009 | Fiji | 14 | 63 | 451 | 2006-07 | Traditional (Burrows, Pruritus) | 40.4 | 0.706 | Western Pacific Region | Oceania | Urban and Rural | Other (Childcare centre, Welfare Home, Clinic/Hospital) | 5825 | Upper Middle Income | Infants |
| 42 | (FJI) Tsoi et al | 2021 | Fiji | 14.00 | 470 | 3351 | 2019 | IACS | 30.7 | 0.746 | Western Pacific Region | Oceania | Urban and Rural | Community | 5968.3 | Upper Middle Income | All ages |
| 43 | (WSM) Taiaroa et al | 2020 | Samoa | 14.4 | 120 | 833 | 2018 | Intergrated Management of Childhood Illeness (IMCI) | 38.7 | 0.716 | Western Pacific Region | Oceania | Rural | School | 4189 | Upper Middle Income | 4-15 years |
| 44 | (SLB) Lake et al | 2021 | Solomon Islands | 15 | 787 | 5239 | 2019 | IACS | 37.1 | 0.567 | Western Pacific Region | Oceania | Rural | Community | 2398.8 | Lower Middle Income | All ages |
| 45 | (MWI) Casas et al | 2021 | Malawi | 15.40 | 2392 | 15487 | 2018 | Traditional (Burrows, Pruritus) | 44.7 | 0.51 | African Region | Africa | Rural | Community | 537.9 | Low Income | All ages |
| 46 | (GMB) Armitage et al | 2019 | Gambia | 15.9 | 229 | 1441 | 2018 | Intergrated Management of Childhood Illeness (IMCI) | 35.9 | 0.495 | African Region | Africa | Urban | Community | 758 | Low Income | <5 years |
| 47 | (AUS) Tasani et al | 2016 | Australia | 16.5 | 84 | 508 | 2016 | Traditional (Burrows, Pruritus) | 33.7 | 0.935 | Western Pacific Region | Oceania | Rural | Community | 49875 | High Income | <13 years |
| 48 | (TLS) Santos et al | 2010 | Timor-Leste | 17.3 | 266 | 1535 | 2007 | Traditional (Burrows, Pruritus) | 27.8 | 0.585 | South-East Asian Region | Asia | Urban and Rural | Community | 533 | Lower Middle Income | 4 months-97 years |
| 49 | (CMR) Kouotou et al | 2016 | Cameroon | 17.80 | 338 | 1902 | 2015 | Traditional (Burrows, Pruritus) | 46.6 | 0.56 | African Region | Africa | Urban | School | 1399.7 | Lower Middle Income | 9-22 years |
| 50 | (BWA) Rainer et al | 2024 | Botswana | 18.2 | 78 | 429 | 2022 | IACS | 53.3 | 0.708 | African Region | Africa | Rural | Community | 7726 | Upper Middle Income | All ages |
| 51 | (FJI) Steer et al | 2009 | Fiji | 18.5 | 640 | 3462 | 2006-07 | Traditional (Burrows, Pruritus) | 40.4 | 0.706 | Western Pacific Region | Oceania | Urban and Rural | School | 5825 | Upper Middle Income | 5-15 years |
| 52 | (SLB) Marks et al | 2020 | Solomon Islands | 18.7 | 261 | 1399 | 2015 | Intergrated Management of Childhood Illeness (IMCI) | 37.1 | 0.559 | Western Pacific Region | Oceania | Rural | Community | 2134 | Lower Middle Income | All ages |
| 53 | (SLB) Mason et al | 2016 | Solomon Islands | 19.2 | 366 | 1908 | 2014 | Traditional (Burrows, Pruritus) | 37.1 | 0.558 | Western Pacific Region | Oceania | Rural | Community | 2235 | Lower Middle Income | All ages |
| 54 | (ETH) Ararsa et al | 2023 | Ethiopia | 19.26 | 88 | 457 | 2021 | IACS | 35 | 0.498 | African Region | Africa | Urban | Community | 925 | Low Income | 5-14 years |
| 55 | (ETH) Melese et al | 2023 | Ethiopia | 21.5 | 203 | 942 | 2021 | Traditional (Burrows, Pruritus) | 35 | 0.498 | African Region | Africa | Rural | Community | 925 | Low Income | All ages |
| 56 | (TLS) Korte et al | 2018 | Timor-Leste | 22.4 | 312 | 1396 | 2016 | Traditional (Burrows, Pruritus) | 28.7 | 0.604 | South-East Asian Region | Asia | Urban and Rural | School | 1348 | Lower Middle Income | 5-24 years |
| 57 | (FJI) Steer et al | 2009 | Fiji | 23 | 105 | 457 | 2006-07 | Traditional (Burrows, Pruritus) | 40.4 | 0.706 | Western Pacific Region | Oceania | Urban and Rural | School | 5825 | Upper Middle Income | 5-15 years |
| 58 | (FJI) Romani et al | 2015 | Fiji | 23.6 | 2564 | 10887 | 2007 | Traditional (Burrows, Pruritus) | 40.4 | 0.706 | Western Pacific Region | Oceania | Urban and Rural | Community | 3793 | Upper Middle Income | All ages |
| 59 | (ETH) Haile et al | 2020 | Ethiopia | 23.8 | 139 | 583 | 2019 | Traditional (Burrows, Pruritus) | 35 | 0.498 | African Region | Africa | Urban and Rural | Community | 840 | Low Income | <15 years |
| 60 | (VUT) Callum et al | 2019 | Vanuatu | 30 | 563 | 1879 | 2019 | Traditional (Burrows, Pruritus) | 32.3 | 0.611 | Western Pacific Region | Oceania | Rural | Community | 3077 | Lower Middle Income | All ages |
| 61 | (MYS) Zayyid et al | 2010 | Malaysia | 31 | 37 | 120 | Not Reported | Traditional (Burrows, Pruritus) | 45.5 | 0.762 | Western Pacific Region | Asia | Urban | Other (Childcare centre, Welfare Home, Clinic/Hospital) | 7167 | Upper Middle Income | 4-18 years |
| 62 | (FJI) Haar et al | 2014 | Fiji | 31.80 | 242 | 760 | 2004 | Traditional (Burrows, Pruritus) | 38.1 | 0.704 | Western Pacific Region | Oceania | Rural | Community | 3124 | Lower Middle Income | All ages |
| 63 | (CMR) Kouotou et al | 2017 | Cameroon | 32.1 | 242 | 755 | 2014 | Traditional (Burrows, Pruritus) | 46.6 | 0.553 | African Region | Africa | Urban | Other (Childcare centre, Welfare Home, Clinic/Hospital) | 1632 | Lower Middle Income | All ages |
| 64 | (TLS) Matthews et al | 2021 | Timor-Leste | 33.4 | 348 | 1043 | 2019 | IACS | 28.7 | 0.614 | South-East Asian Region | Asia | Urban and Rural | School | 1584 | Lower Middle Income | <19 years |
| 65 | (ETH) Enbiale et al | 2018 | Ethiopia | 33.5 | 379000 | 1125770 | 2015 | Traditional (Burrows, Pruritus) | 35 | 0.46 | African Region | Africa | Urban and Rural | Community | 630 | Low Income | All ages |
| 66 | (TLS) Tsoi et al | 2021 | Timor-Leste | 33.80 | 321 | 951 | 2019 | IACS | 28.7 | 0.614 | South-East Asian Region | Asia | Urban and Rural | School | 1584 | Lower Middle Income | <19 years |
| 67 | (BAN) Hasan et al | 2024 | Bangladesh | 33.9 | 317 | 935 | 2023 | IACS | 33.4 | 0.67 | South-East Asian Region | Asia | Rural | Other (Childcare centre, Welfare Home, Clinic/Hospital) | 2529 | Lower Middle Income | 3-18 years |
| 68 | (FJI) Romani et al | 2017 | Fiji | 36.4 | 746 | 2051 | 2012-2013 | Intergrated Management of Childhood Illeness (IMCI) | 36.7 | 0.726 | Western Pacific Region | Oceania | Rural | Community | 4586 | Upper Middle Income | All ages |
| 69 | (SLB) Osti et al | 2019 | Solomon Islands | 54.3 | 176 | 324 | 2018 | IACS | 37.1 | 0.566 | Western Pacific Region | Oceania | Urban and Rural | School | 2451 | Lower Middle Income | 4-15 years |
| 70 | (GHA) Amoako et al | 2020 | Ghana | 71 | 200 | 283 | 2019 | IACS | 43.5 | 0.631 | African Region | Africa | Urban and Rural | Community | 2167 | Lower Middle Income | All ages |

**Figure S5: Leave-one out forest plot**

| (a) Studies sorted by proportion | (b) Studies sorted by heterogeneity |
| --- | --- |
| 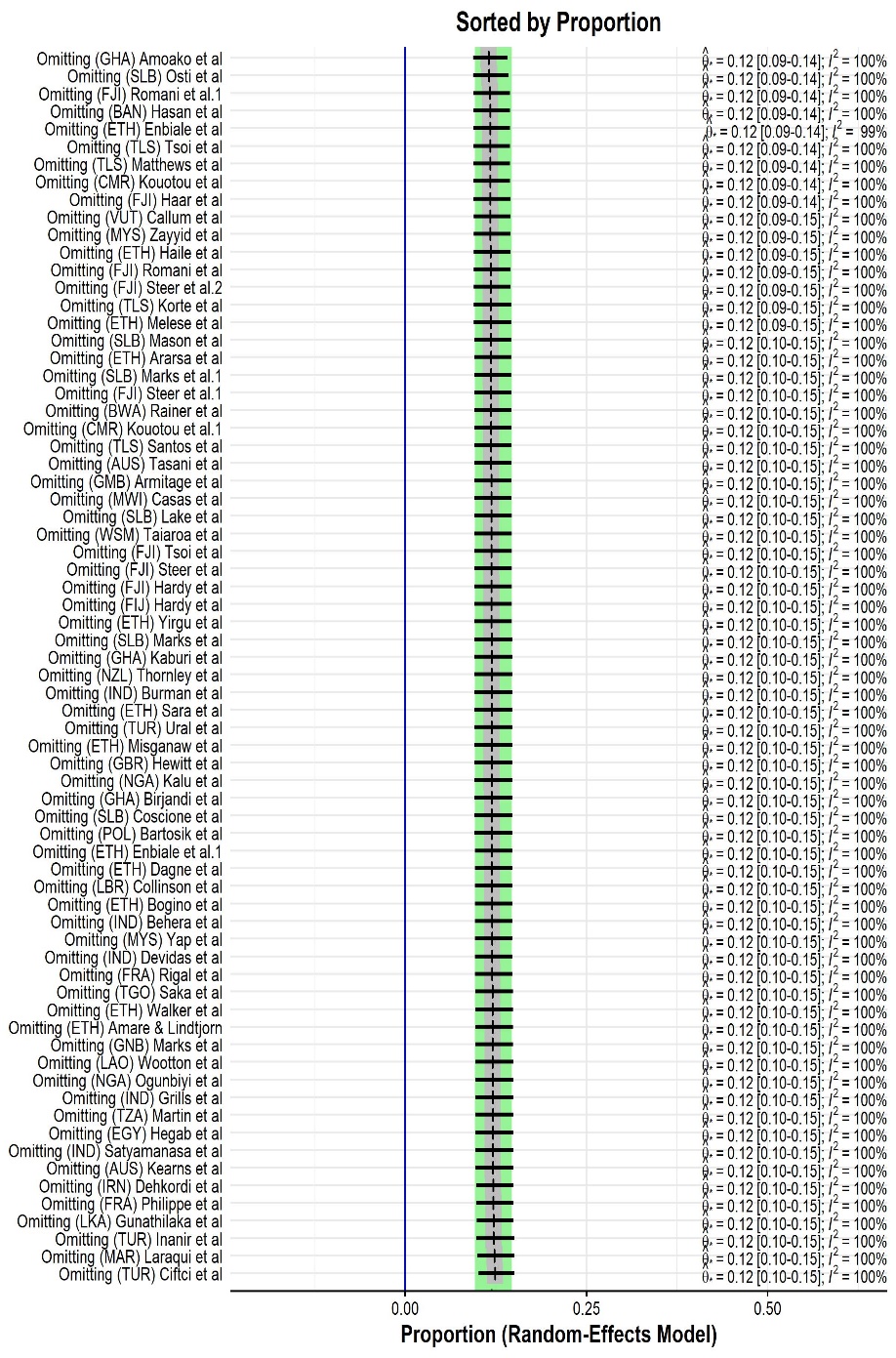 | 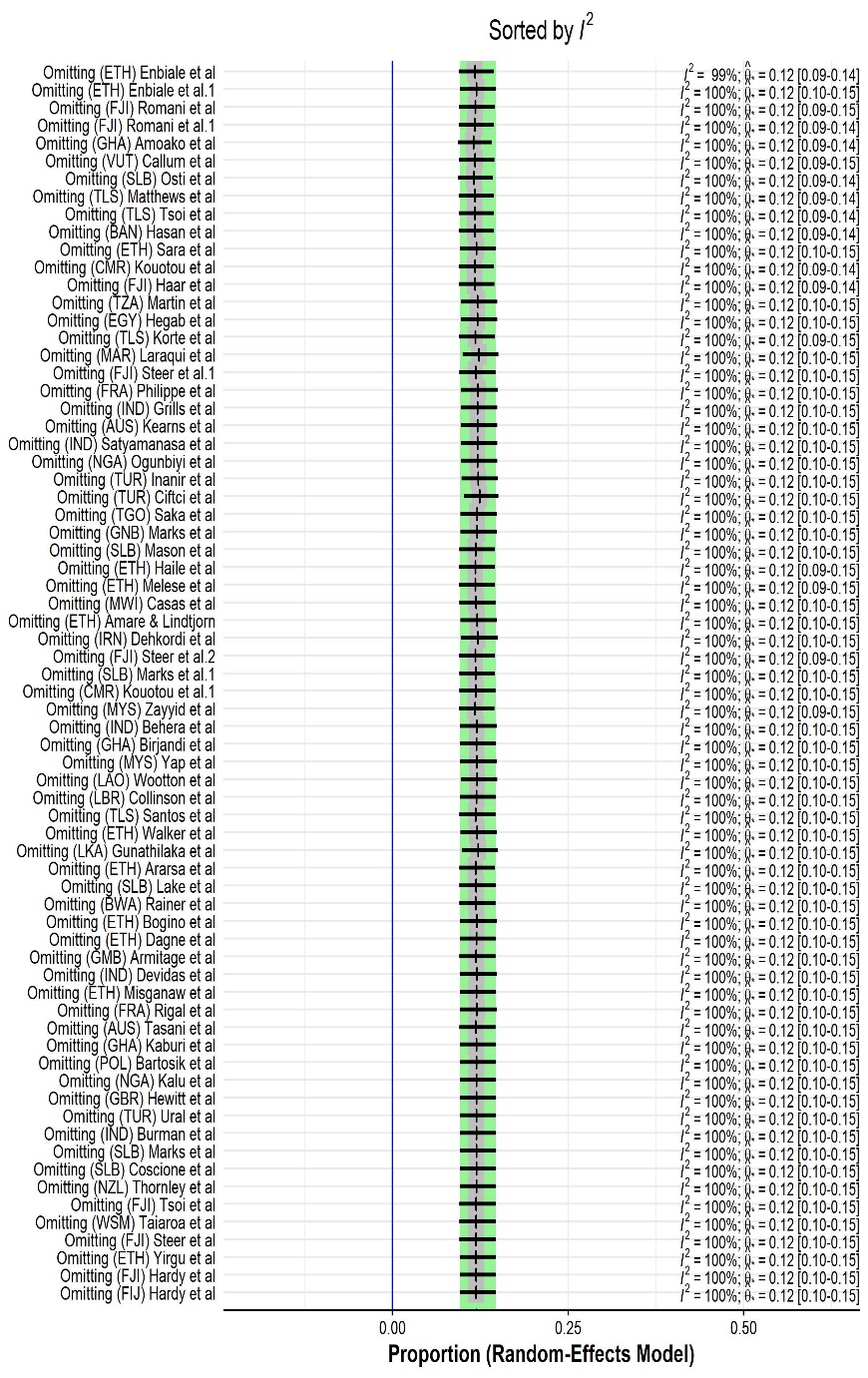 |

**Figure S6: Influence Analysis**

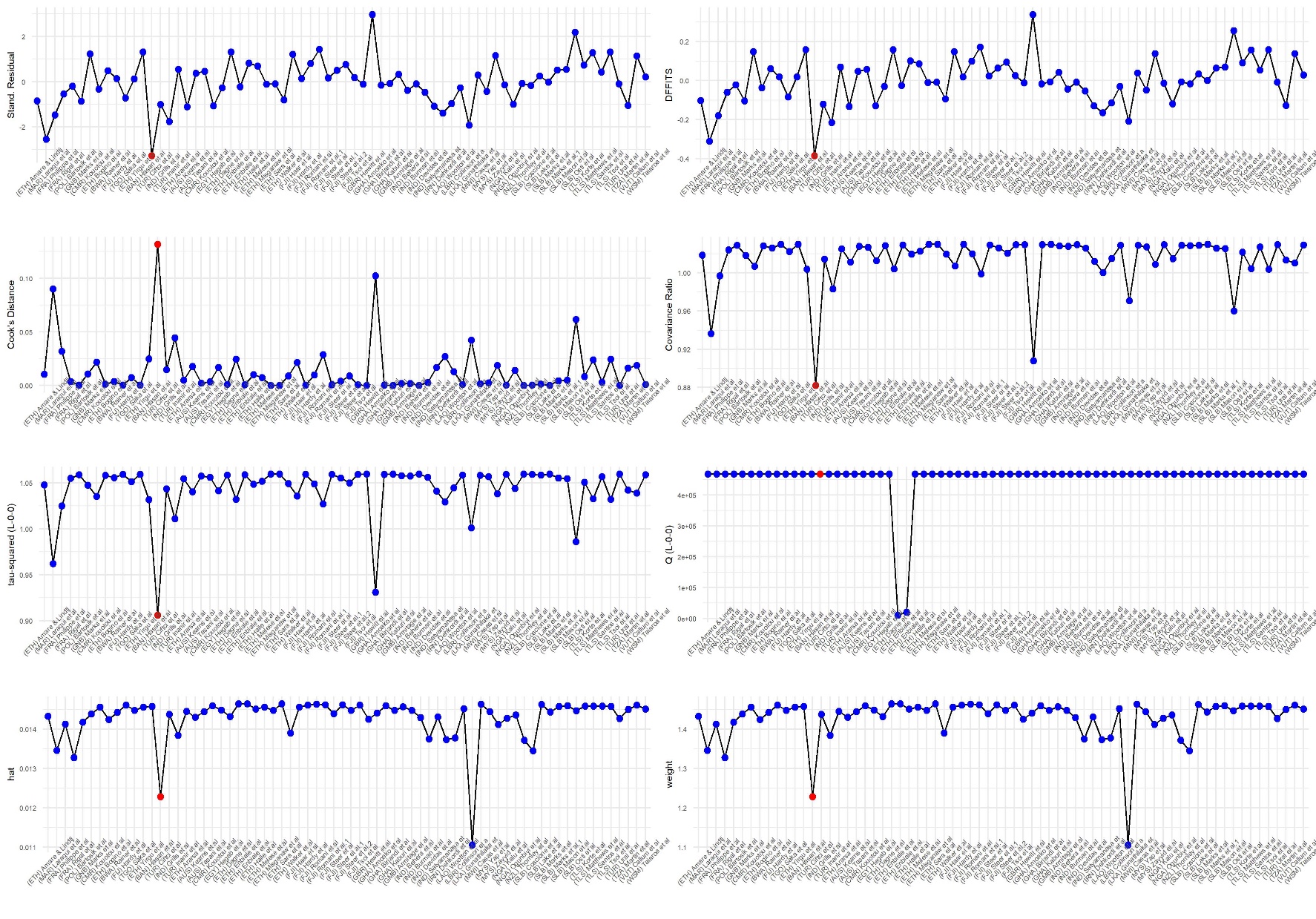

**Figure S7: Baujat Plot showing the outlier studies according to heterogeneity.**

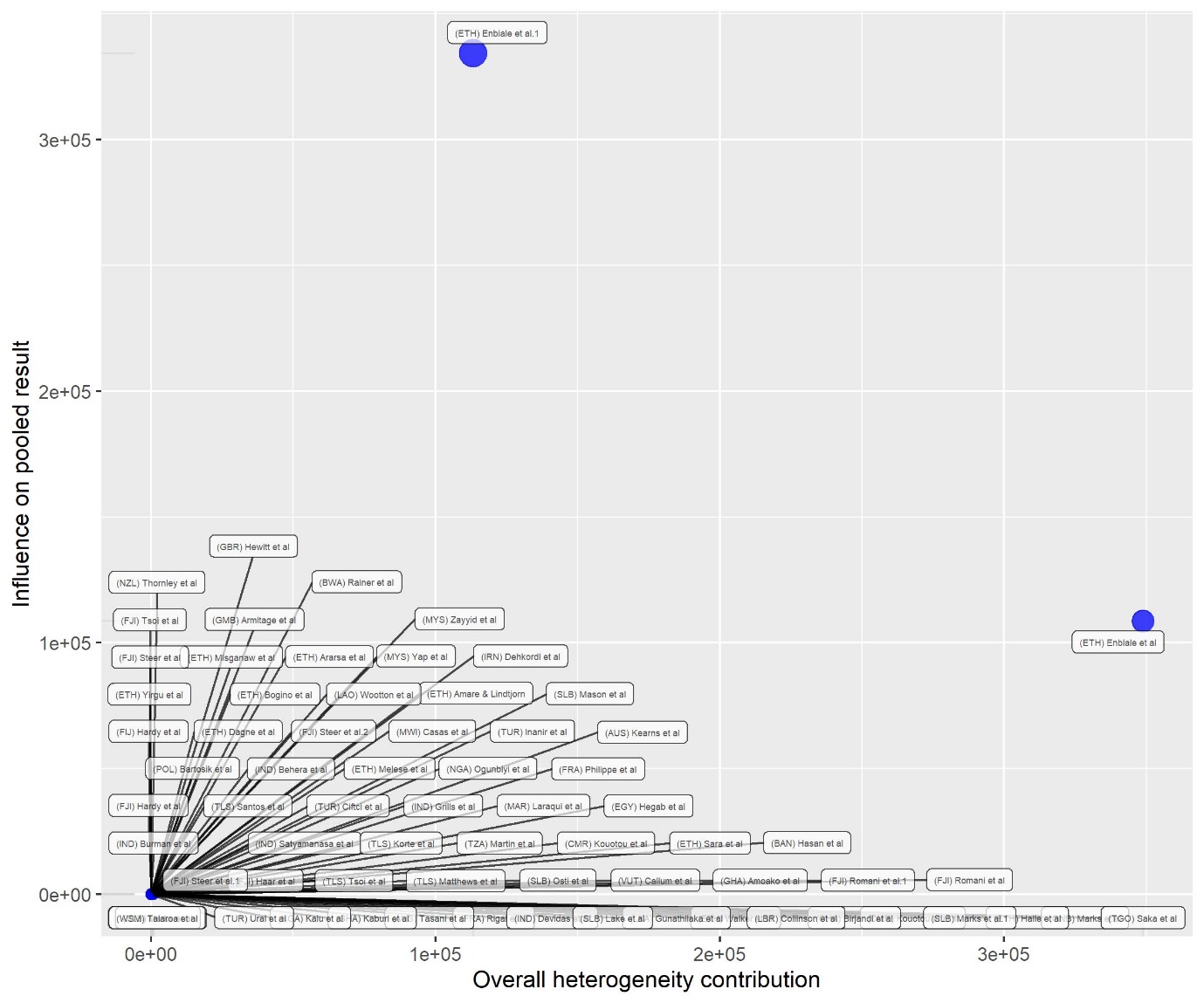

**Table S8: Meta-regression of scabies prevalence and socioeconomic factors**

|  | *β* coefficient | standard error | *p*-value | Lower 95% CI | Upper 95% CI |
| --- | --- | --- | --- | --- | --- |
| Model 1 | -0.17 | 0.49 | 0.72 | -1.14 | 0.79 |
| Model 2 | -0.04 | 0.08 | 0.67 | -0.19 | 0.12 |
| Model 3 | -0.10 | 0.15 | 0.50 | -0.40 | 0.19 |

Model 1: logit GINI vs logit prevalence; Model 2: log GDP vs log prevalence; Model 3 logit HDI vs logit prevalence

**Table S9: Risk of publication bias.**

|  | Harbord Test | | | Peters Test | | |
| --- | --- | --- | --- | --- | --- | --- |
|  | Bias | t | p-value | Bias | t | p-value |
| Family size | 0.48 | 0.24 | 0.82 | 80.31 | 0.63 | 0.56 |
| Share clothes | 4.95 | 2.67 | 0.05 | 465.7 | 1.33 | 0.24 |
| Bed sharing | -2.22 | -0.77 | 0.48 | 22.12 | 0.22 | 0.84 |
| Gender | 0.91 | 0.97 | 0.34 | 63.29 | 0.68 | 0.50 |
| Location | -0.65 | -0.12 | 0.91 | 169.74 | 0.19 | 0.86 |
| Frequency of bathing | 5.18 | 1.67 | 0.17 | 84.58 | 0.20 | 0.85 |
| Source of water | 5.53 | 0.72 | 0.54 | 378.08 | 1.62 | 0.24 |
| Contact with itchy person | 5.61 | 1.02 | 0.36 | 92.45 | 0.15 | 0.88 |
| Presence of pets | -0.92 | -0.15 | 0.89 | 268.4 | 0.42 | 0.71 |
| Soap use | 1.55 | 0.90 | 0.41 | 1.55 | 0.90 | 0.41 |
